# Correlation between universal BCG vaccination policy and reduced mortality for COVID-19

**DOI:** 10.1101/2020.03.24.20042937

**Authors:** Aaron Miller, Mac Josh Reandelar, Kimberly Fasciglione, Violeta Roumenova, Yan Li, Gonzalo H. Otazu

**Affiliations:** Department of Biomedical Sciences, NYIT College of Osteopathic Medicine, New York Institute of Technology, Old Westbury, New York, USA

## Abstract

COVID-19 has spread to most countries in the world. Puzzlingly, the impact of the disease varies in different countries. This variation is attributed to differences in cultural norms, mitigation efforts, and health infrastructure. Here, we propose that national differences in COVID-19 impact could be partially explained by different national policies with respect to Bacillus Calmette-Guérin (BCG) vaccination. BCG vaccination has been reported to offer broad protection from other respiratory infections besides tuberculosis. We compared BCG vaccination policies with the morbidity and mortality for COVID-19 for middle-high and high-income countries. We found that countries without universal policies of BCG vaccination (Italy, the Netherlands, USA) have been more severely affected compared to countries with universal and long-standing BCG policies. The difference cannot be accounted for by differences in disease onset, adoption of early social distancing policies, state of health services, nor income level. Reduced mortality suggests BCG vaccination could be a potential new tool in the fight against COVID-19.

## Introduction

The COVID-19 pandemic originated in China and has quickly spread across the world, affecting nearly every country. However, there are some striking differences in how COVID-19 is behaving in different countries. For instance, in Italy there has been strong curtailing of social interactions but COVID-19 mortality is high (29315 deaths as of May 5^th^, 2020). In contrast, Japan had some of the earlier cases, but the mortality is low (556 deaths as of May 5^th^, 20202) despite not having adopted some of the more restrictive social isolation measurements (“Japan Was Expecting a Coronavirus Explosion. Where Is It? | The Japan Times” n.d.). These puzzling differences have been adjudicated to different cultural norms as well as differences in medical care standards. In addition, it has been proposed that the reduced mortality in children might be due to recent vaccination(Cao et al. 2020). Here we propose that the country-by-country difference in COVID-19 mortality can be partially explained by national policies on Bacillus Calmette-Guérin (BCG) vaccination.

BCG is a live attenuated strain derived from an isolate of *Mycobacterium bovis* used widely across the world as a vaccine for Tuberculosis (TB), with many nations, including Japan and China, having a universal BCG vaccination policy in newborns(“WHO | Tuberculosis” n.d.). Other countries such as Spain, France, and Switzerland, have discontinued their universal vaccine policies due to comparatively low risk for developing *M. bovis* infections as well as the proven variable effectiveness in preventing adult TB. Countries such as the United States, Italy, and the Netherlands, have yet to adopt universal vaccine policies for similar reasons.

Several vaccines including the BCG vaccination have been shown to produce positive “heterologous” or non-specific immune effects leading to improved response against other non-mycobacterial pathogens. For instance, in severe combined immunodeficiency (SCID) mice, which lack functional B- and T-cells, BCG vaccination protected against a secondary non-mycobacterial challenge, demonstrating the ability of innate immune cells to mount a non-specific ‘memory-like’ response(Kleinnijenhuis et al. 2014). This phenomenon was named ‘trained immunity’ and is proposed to be caused by metabolic and epigenetic changes leading to the promotion of genetic regions encoding for pro-inflammatory cytokines(Netea et al. 2016). BCG vaccination significantly increases the secretion of pro-inflammatory cytokines, specifically IL-1B, which has been shown to play a vital role in antiviral immunity(Kleinnijenhuis et al. 2014). Additionally, a study in Guinea-Bissau found that children vaccinated with BCG were observed to have a 50% reduction in overall mortality, which was attributed to the vaccine’s effect on reducing respiratory infections and sepsis (Kristensen, Aaby, and Jensen 2000) i.

Reports on the duration of BCG vaccination protection against TB vary between 10 years(Sterne, Rodrigues, and Guedes 1998) and 60 years (Aronson et al. 2004). BCG protection for conditions other than TB has also shown long-lasting protection: a 60-year follow up of a clinical trial suggested that individuals who received the BCG vaccination during early childhood subsequently demonstrated a 2.5 fold reduction of lung cancer development (Usher et al. 2019). Whether the mechanism responsible for the BCG vaccination’s long-standing effectiveness against mycobacterial pathogens carries over as a means of protection against the novel COVID-19 remains unknown, but the potential opens up an interesting area for exploration.

Given our current understanding of the BCG vaccine’s nonspecific immunotherapeutic mechanisms and by analyzing current epidemiological data, this investigation aims to identify a possible correlation between the existence of universal BCG vaccine policies and mortality associated to COVID-19 infections all over the world.

## Methods

We collected the BCG vaccination policies across countries from the BCG World Atlas (Zwerling et al. 2011), available from http://www.bcgatlas.org/. We complemented the database with respect to the dates of initiation of BCG vaccination. The additional references are in the adjunct table. Data on COVID-19 cases and death per country were obtained from https://google.org/crisisresponse/covid19-map on May 5th, 2020. We included in the analysis only countries with more than 1 million inhabitants. The mortality rate might be influenced by multiple factors including a country’s economics and the age distribution of the population. To account for economical differences, we classified countries according to their GNI per capita in 2018 using the World Bank data (https://datahelpdesk.worldbank.org/knowledgebase/articles/906519-world-bank-country-and-lending-groups). Countries were divided into three categories: low income (L) with an annual income of 1,025 dollars or less, lower-middle income with an income between 1,026 and 3,995 dollars, and middle-high and high-income countries, which included countries with annual incomes over 3,996 dollars.

COVID-19 mortality increases steeply with age(Zhou et al. 2020). Universal vaccination policies are more common in developing countries which tend to have younger populations. A younger population would show spurious reduced mortality for COVID-19. To control for this variable, we did not use the total number of inhabitants per country to calculate the mortality rate, but we considered the susceptible population as the inhabitants over 65 years of age per country(Shet et al. n.d.). Medians between populations were compared using the Wilcoxon rank sum test. Multivariate linear analyses were done using the function *fitlm* in Matlab (R2017b). The Matlab script is available as supplementary material.

## Results

Initially, we compared countries that never had in place a universal BCG vaccination policy (Italy, USA, Lebanon, Nederland, and Belgium), with countries that have a current universal BCG vaccination policy. Low-income countries report a median of 12.6 deaths per million people over 65 years of age. In fact, 23% (4/17) of these countries reported zero deaths attributed to COVID-19, consistent with the hypothesis for a protective role of BCG vaccination. This effect, however, could be attributed to significantly lower testing rates in low income countries (Spearnman correlation between median income and tests per million inhabitants, rho=0.8083,p<1e-6, n=101 countries). Therefore, we will only consider middle-high and high-income countries for our analysis, which should compare countries with similar levels of medical care standards, sanitation, and testing levels for COVID-19. Middle high and high income countries with universal policies have a high BCG coverage, with a median of 97% with the 10 percentile being 87% and the 90 percentile being 99%.

Middle-high and high-income countries that have a current universal BCG policy (55 countries) had a median of 84.5 deaths per million people over 65 years of age (see **Figure 1**). In contrast, middle-high and high-income countries that never had a universal BCG policy (5 countries) had a higher mortality rate, with a median of 1583.2 deaths per million people over 65 years of age. This difference between the median number of deaths was significant (p=0.008, Wilcoxon rank sum test).

**Figure 1:**
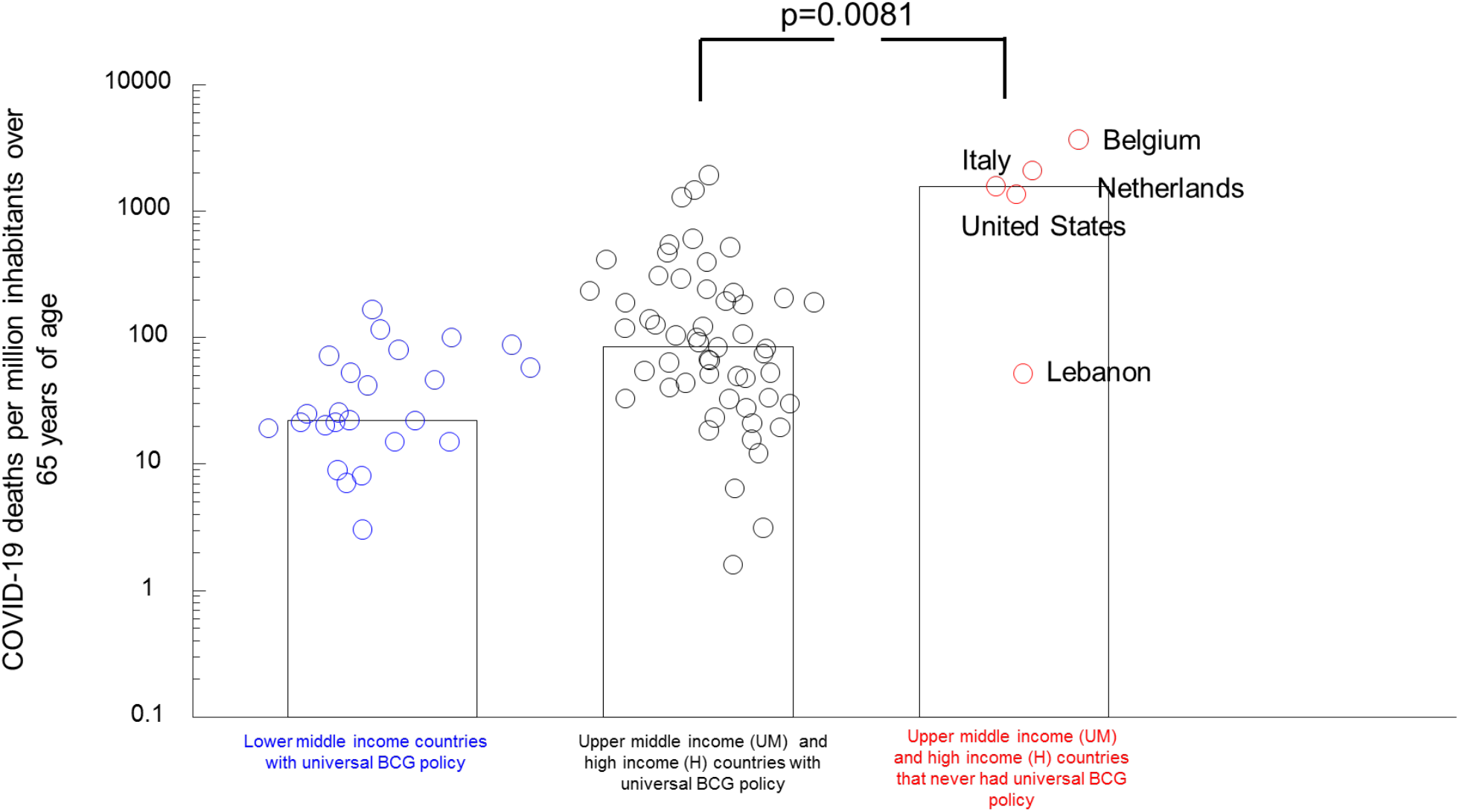
Higher death rates were presented in countries that never implemented a universal BCG vaccination policy. Bars indicate the median value of a given group of countries.

An alternative explanation for the reduced number of deaths in countries with universal BCG vaccination policies could be that COVID-19 arrived late in these countries and that they are in the earlier stages of the epidemic. To evaluate for the significance of a possible confounding effect of epidemic stage, we plotted the number of deaths per million inhabitants over 65 years of age versus the number of days since the first reported case of COVID-19 per country (see **Figure 2**). As expected, there is a general increase in the number of reported deaths (as of 05/05/20) as more days pass since a country’s first reported case. Additionally, middle-high and high-income countries *with* universal BCG vaccination policies that saw larger case volumes relatively early have a median of 105.1 deaths per million people over 65 years of age. This is significantly lower (p= 0.0059, Wilcoxon rank sum test) than the median of 1583.2 for countries in the same economic category that never implemented a universal policy and were affected at comparable times or even later (with at least 68 days passing since the first reported case). Therefore, the increased mortality for countries that never implemented a universal BCG vaccination policy cannot be explained by an earlier epidemic onset.

**Figure 2:**
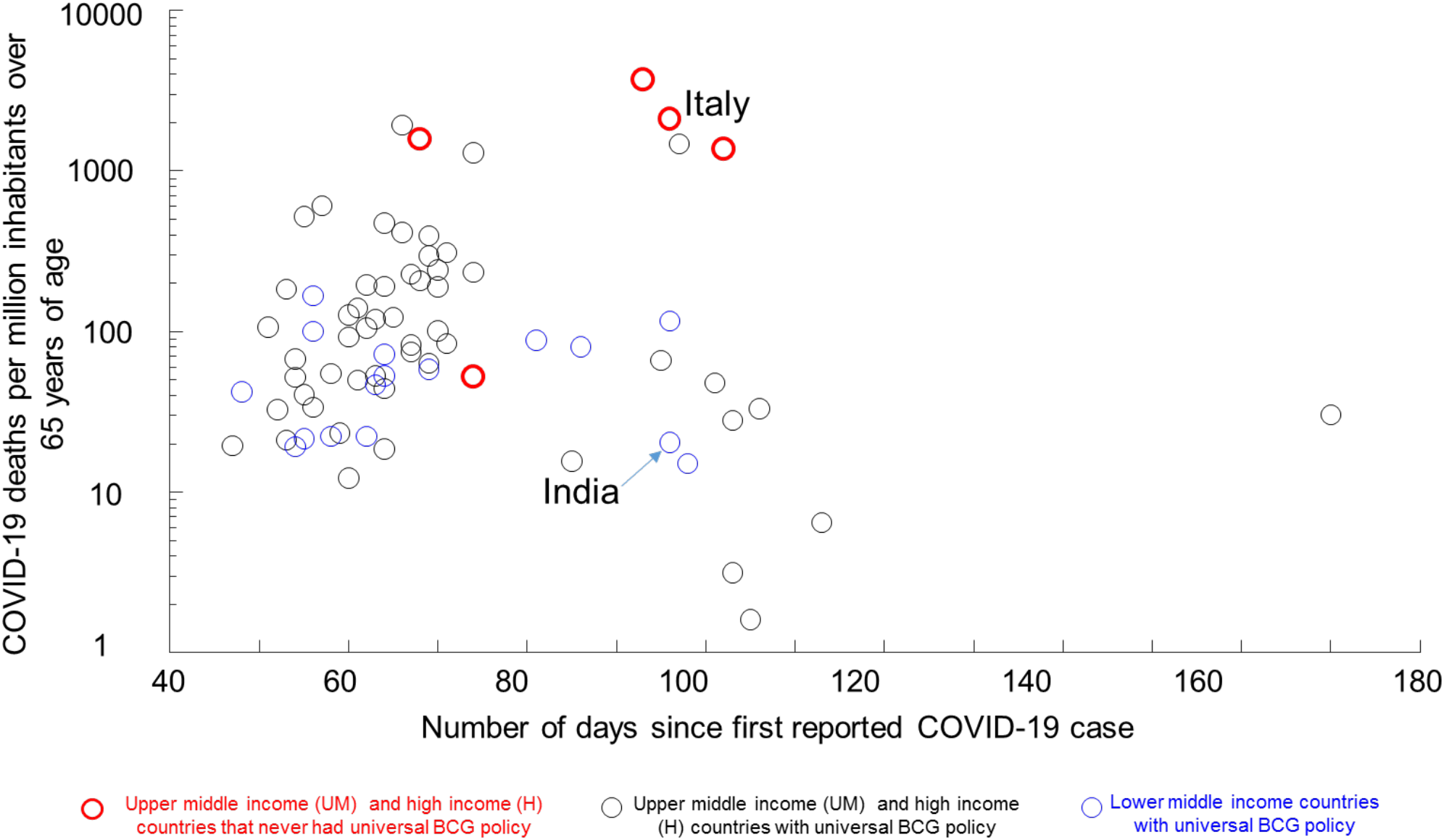
Date of onset of the epidemic does not explain higher number of deaths in countries that did not establish a universal BCG policy.

Although we have stratified the countries and compared only those with middle-high and high-income levels that should be relatively homogeneous, differences in the strength of the health system, poverty rates, and quality of public health policy in response to COVID-19 could still be residual confounders that explain the observed increased death rates in countries that never implemented a universal BCG vaccination policy. In order to address these confounders directly, we used a multivariate linear regression analysis (see Table 1) on the middle-high and high-income countries (55 with BCG policy+6 that never had a policy). We included the days since the first reported case as covariates to account for differences in the stage of the epidemic in a country. We included two variables that accounted for the quality of the public health policy with respect to COVID-19. One was the number of tests performed per million people in a country which would correlate with the strength of a country’s response. The other was the speed of a country’s reaction to the pandemic which we quantified as the number of days between the first detected case and schools closing. We used the median life expectancy in a country as a surrogate for the strength of the health system. Although the countries that we considered were middle-high and high-income countries, we also included the median income per country. Although median income might fail to capture the degree of resources assigned to effective COVID-19 mitigation measures, it would serve as an indicator of possible measures that could be taken. Finally, we included the BCG vaccination policy as a binary variable.

**Table 1.**
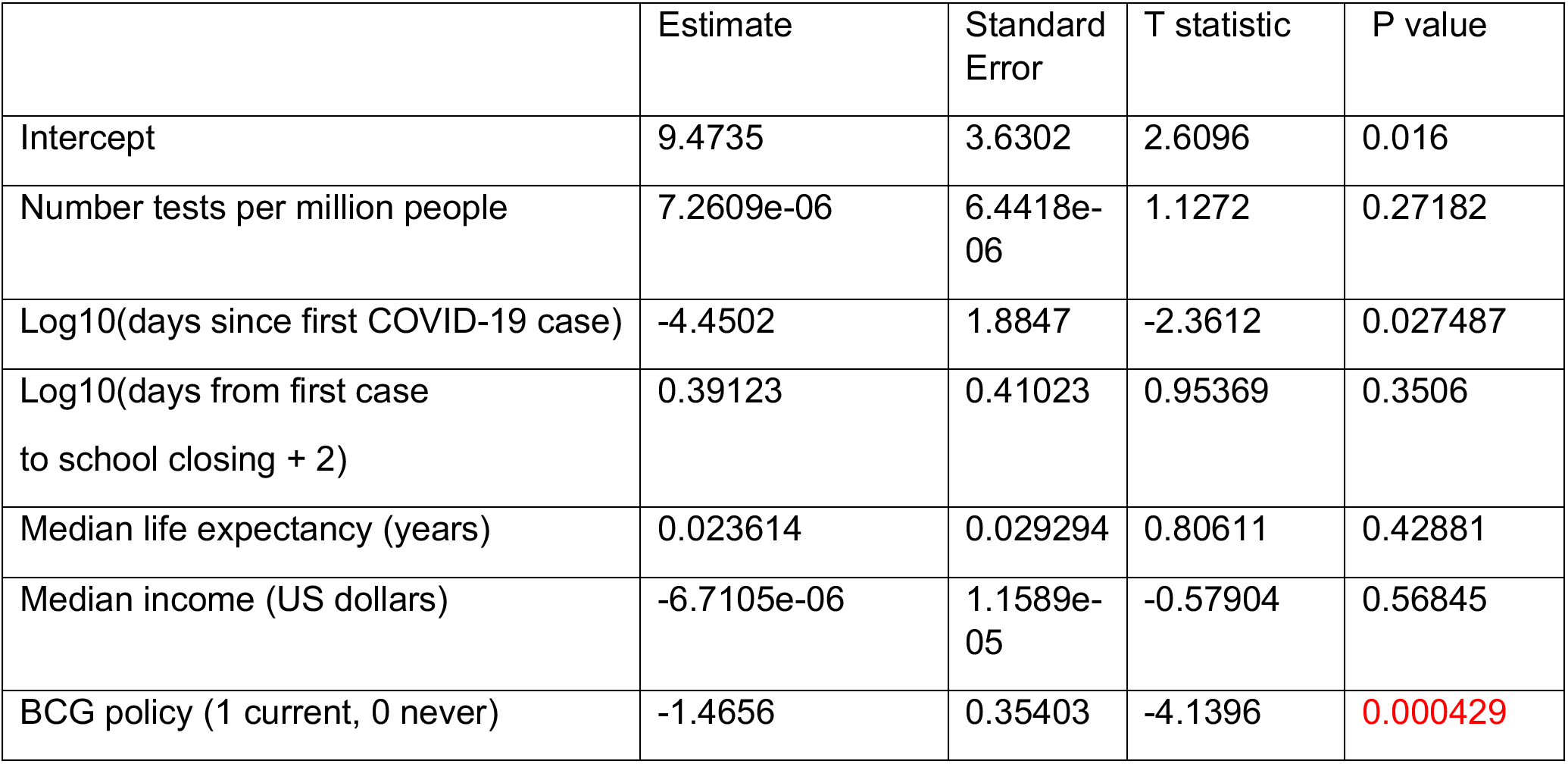
Multivariate analysis of log10(deaths per million inhabitants over 65 years of age). Number of observations: 29, Error degrees of freedom: 22 Root Mean Squared Error: 0.494 R-squared: 0.599, Adjusted R-Squared 0.49 F-statistic vs. constant model: 5.48, p-value = 0.00134

|  | Estimate | Standard Error | T statistic | P value |
| --- | --- | --- | --- | --- |
| Intercept | 9.4735 | 3.6302 | 2.6096 | 0.016 |
| Number tests per million people | 7.2609e-06 | 6.4418e-06 | 1.1272 | 0.27182 |
| Log10(days since first COVID-19 case) | -4.4502 | 1.8847 | -2.3612 | 0.027487 |
| Log10(days from first case to school closing + 2) | 0.39123 | 0.41023 | 0.95369 | 0.3506 |
| Median life expectancy (years) | 0.023614 | 0.029294 | 0.80611 | 0.42881 |
| Median income (US dollars) | -6.7105e-06 | 1.1589e-05 | -0.57904 | 0.56845 |
| BCG policy (1 current, 0 never) | -1.4656 | 0.35403 | -4.1396 | 0.000429 |

Consistent with our previous analysis, we found that the only significant covariate (p=0.000429) was the presence of universal BCG vaccination policy in reducing the mortality of COVID-19. None of the other factors reached significance (p>0.01). However, there still might be other factors (genetic or environmental) that might be producing the correlation between BCG vaccination policy and reduced deaths due to COVID-19.

The number of deaths per million people over the age of 65 years did not correlate with the number of tests performed (p=0.27182), indicating that the number of deaths was a robust indicator that did not depend on a country’s testing response. Nevertheless, we have used data from COVID-19 attributed deaths as reported by each country. There still might be systematic differences across countries on whether a death is attributed to COVID-19. For instance, if countries with universal BCG vaccination would tend to assign COVID-19 deaths to other causes (atypical pneumonia), that could create a spurious correlation between BCG vaccination and reduced mortality. In order to address this concern, we used a smaller dataset of European countries (https://www.euromomo.eu/) that calculated excess deaths in 2020. We extracted the data from weeks 12 to 17 of the year 2020 (see **Figure 3**) when the epidemic was peaking in Europe. Countries with universal BCG vaccination policy had median excess deaths per week of 0.42 (n=5), whereas countries without universal BCG vaccination policies had significantly higher median excess deaths of 16.02 per week (n=3, p=0.035, Wilcoxon rank sum test). Albeit a small sample, this independent measurement supports the idea that the reported correlation between a country’s BCG vaccination and reduced mortality due to COVID-19 is not an artifact of national differences in COVID-19 death assignation.

**Figure 3:**
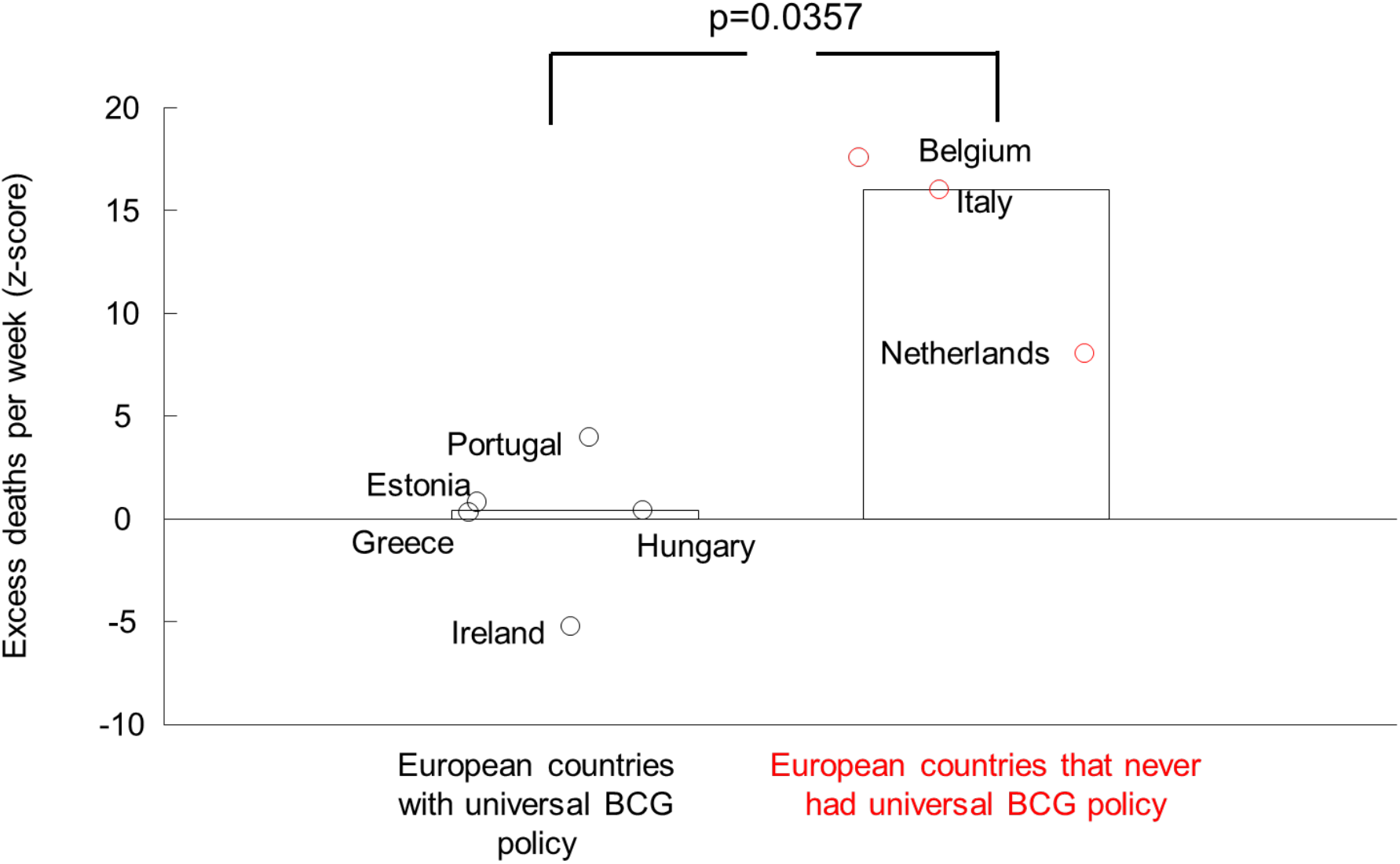
Higher excess deaths were presented in European countries that never implemented a universal BCG vaccination policy. Bars indicate the median value of a given group of countries.

A possible mechanism for a putative protective effect of BCG in a population might include direct protection in the elderly population, which is the most affected group(Zhou et al. 2020), as well as herd immunity acting over the whole population, both young and old. Using data from March 21^st^, 2020(Miller et al. 2020), we reported a negative correlation between the year start of the vaccination policy and mortality both in middle high and high income countries with current policies, as well as countries that had stopped their policies. This was consistent with BCG vaccination producing a long-lasting protection to the elderly population from BCG vaccination received in childhood. However, using the current data from May 5^th^, there is no significant correlation between start year of the vaccination and mortality per million people over 65 years of age (see **Fig. 4**). We do not know if the early reported correlation was spuriously produced by the evolving nature of the pandemic, or if earlier response to the disease are less affected by a country’s pandemic mitigation responses and depend more on intrinsic immunity in the elderly population to COVID-19. This would be consistent with a recent pre-print report (Berg et al. 2020) suggesting that BCG vaccination policy slows down the spread of COVID-19, making it easier to observe differences in the protective effect of BCG earlier in the epidemic.

**Figure 4:**
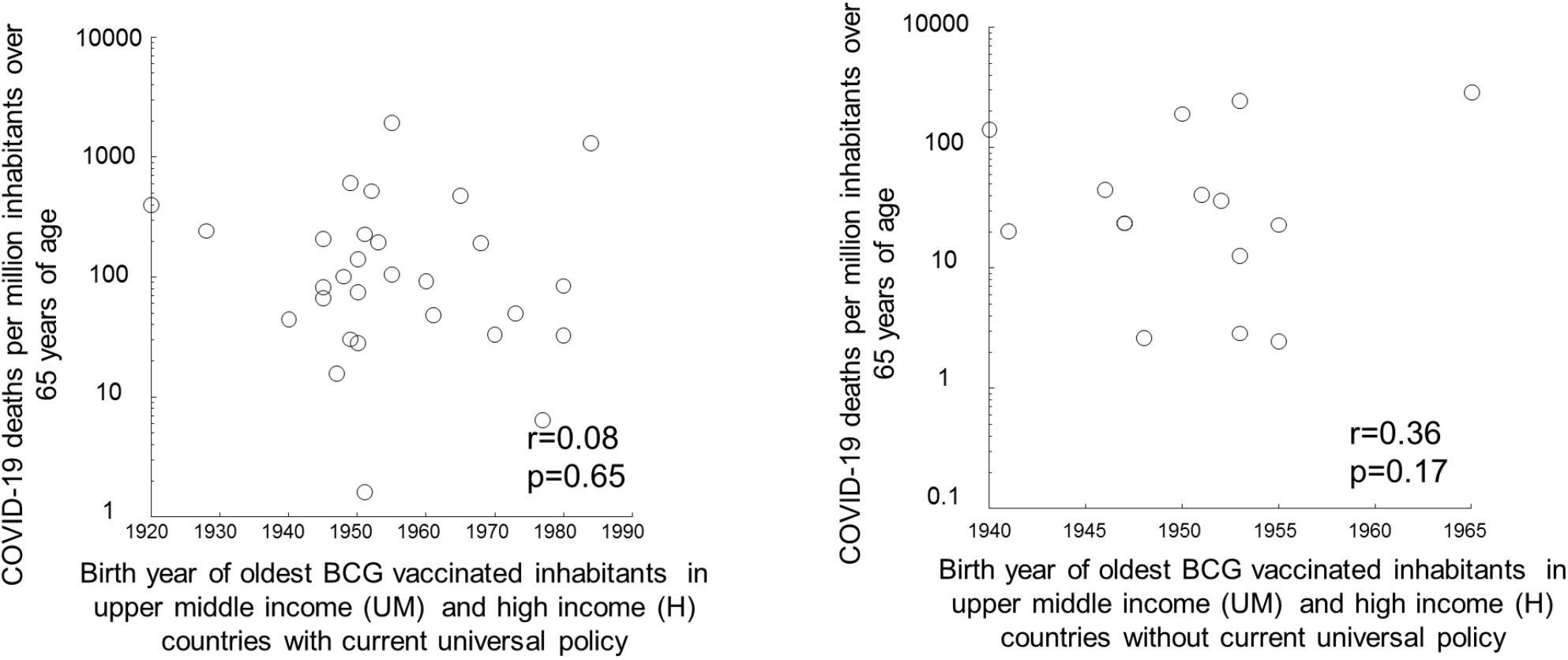
Earlier date of the start of vaccination does not correlate with reduced mortality. Left panel correspond to upper middle income and high income countries with current universal BCG vaccination policy. The right panel corresponds to countries that do not have a current universal vaccination policy.

A second possibility for a putative protective effect of BCG for COVID-19 could be a reduction in the spread of the disease, acting across the population. This would be reflected as a reduction in the number of COVID-19 reported cases. Consistent with this hypothesis, the countries with low-income levels that have a universal policy (16 countries) reported a smaller number of cases of COVID-19 per million inhabitants with a median of 8.85. However, the issue of underreporting might be more critical for estimating the number of cases and we have excluded the low-income countries from further analysis. Middle-high and high-income countries that have a current universal BCG vaccination policy (55 countries) had a median of 313.6 cases per million inhabitants (see **Figure 5**). Consistent with the role of BCG in slowing spread of COVID-19; middle-high and high-income countries that never had a universal BCG policy (5 countries) had about 10 times the number of cases, with a median of 3522.0 cases per million inhabitants. This difference in the medians between countries was significant (p=0.0283, Wilcoxon rank sum test), suggesting that broad BCG vaccination along with other measures could help slow the spread of COVID-19. However, multivariate analysis of the number of cases (see **Table 2**) revealed that although the BCG vaccination policy had a negative correlation with the log_10_ number of COVID-19 reported cases, the factor did not reach significance (p=0.12). The most significant factor (p=0.00599) in determining the number of cases per million inhabitants was the number of tests performed per million inhabitants. Conclusions based on the number of deaths per million inhabitants over 65 years of age are more robust than conclusions based on the number of cases per million inhabitants as they do not depend on the number of tests performed.

**Figure 5:**
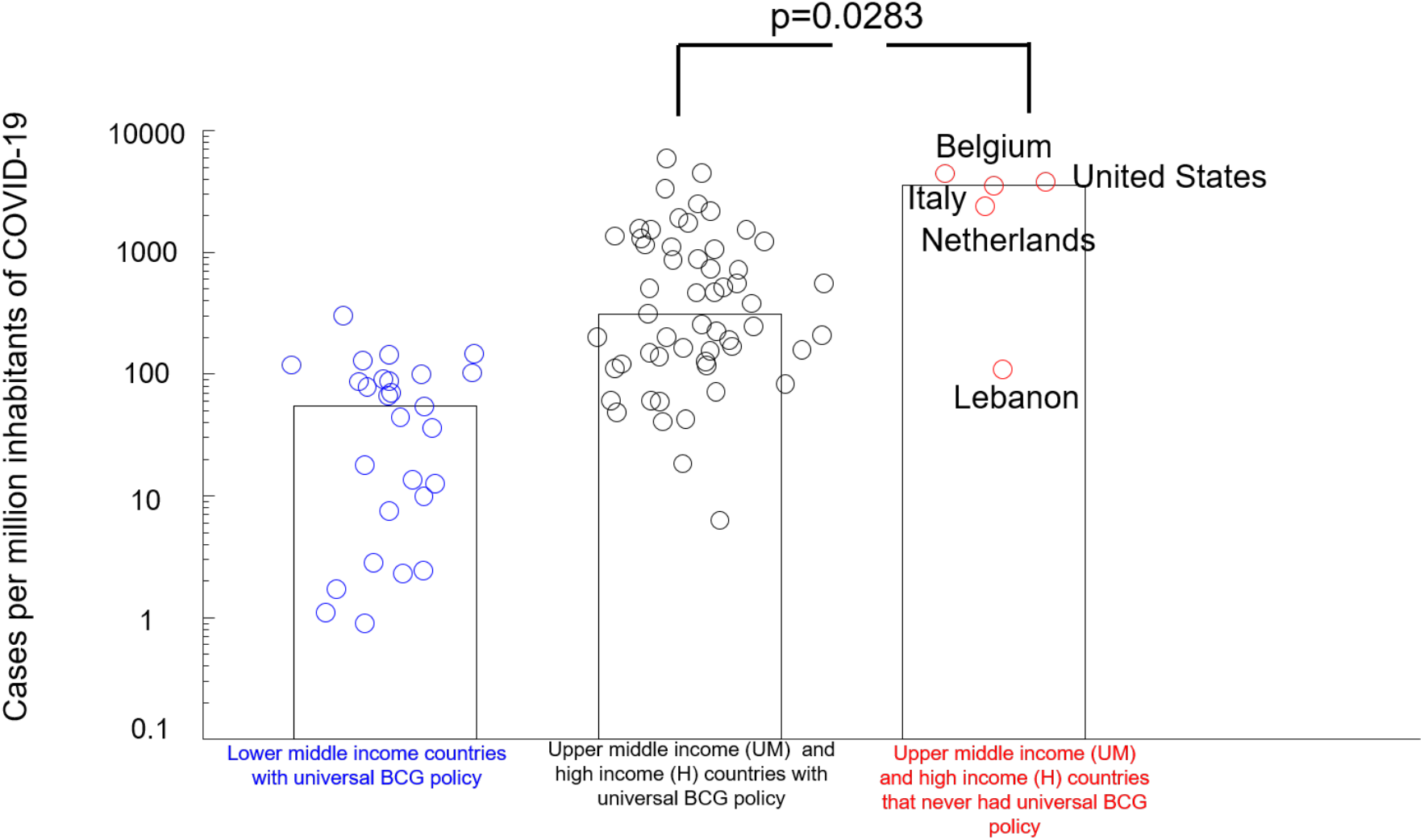
Higher number of COVID-19 cases were presented in countries that never implemented a universal BCG vaccination policy. Bars indicate the median value of a given group of countries.

**Table 2.** Multivariate analysis of log10(number of cases per million inhabitants). Number of observations: 29, Error degrees of freedom: 22 Root Mean Squared Error: 0.368 R-squared: 0.678, Adjusted R-Squared 0.591 F-statistic vs. constant model: 7.73, p-value = 0.000148

|  | Estimate | Standard Error | T statistic | P value |
| --- | --- | --- | --- | --- |
| Intercept | 5.9208 | 2.7057 | 2.1883 | 0.039559 |
| Number tests per million people | 1.46e-05 | 4.8013e-06 | 3.0408 | 0.005999 |
| Log10(days since first case) | -2.8594 | 1.4047 | -2.0356 | 0.054015 |
| Log10(days from first case<br>To school closing + 2) | 0.3817 | 0.30576 | 1.2484 | 0.22502 |
| Median life expectancy (years) | 0.02121 | 0.021834 | 0.97144 | 0.34189 |
| Median income(US dollars) | 1.1029e-05 | 8.6378e-06 | 1.2769 | 0.21497 |
| BCG policy (1 current, 0 never) | -0.42549 | 0.26387 | -1.6125 | 0.12111 |

## Discussion

We have shown that differences in mortality produced by COVID-19 across countries are correlated with a country’s BCG vaccination policy. We centered our analysis in middle-high and high-income countries because they constitute a relatively homogenous group of countries and under the assumption that reported COVID-19 statistics from these countries would be more reliable than statistics from less wealthy countries. We performed multivariate analysis on those countries that controlled for the age distribution of the population, income per capita, stage of the epidemic of a country, and quality of the medical care. We also included as covariates factors related to a country’s response to COVID-19. These other factors did not explain the lower mortality in countries with mandatory BCG vaccination. The correlation was confirmed using an independent measure, excess deaths, from a smaller dataset from Europe.

Although these confounders did not explain the observed trend, other unaddressed factors could be the causal link between BCG vaccination and reduced mortality. This includes complex factors such as the genetic makeup of the population, as well as environmental factors. Therefore, our correlational study should not be the basis for changes in clinical practice nor public health policies. Those changes, if warranted, should be based on the results of well-controlled clinical trials that could demonstrate a causal relationship between BCG vaccination and COVID-19 outcomes. At this time, there are currently 14 clinical trials registered at the NIH (https://clinicaltrials.gov/ct2/results?cond=covid&term=bcg&cntry=&state=&city=&dist=) exploring this possibility.

We did not find a correlation between early implementation of a universal vaccination policy and reduced mortality in middle-high and-high income countries. We did not include in our analysis middle-low or low-income countries because our database might not capture the exact BCG vaccination policies of former European colonies in Asia and Africa during colonial times. For instance, BCG Atlas lists for Vietnam a start date of 1985 for universal BCG vaccination, but there are reports of widespread use of BCG vaccination during the French colonial period (Monnais 2006). Interestingly, Vietnam with a population of 95 million has reported zero fatalities due to COVID-19.

Although COVID-19 deaths are concentrated in the elderly population(Zhou et al. 2020), recent BCG vaccination in children might play a role in reducing mortality by decreasing the transmission of the disease to the vulnerable population. Asymptomatic children in Germany(Jones et al., n.d.), where BCG is not applied to children since 1998, had viral loads that were as high as adults, which suggests that unvaccinated children may be as infectious as adults. A reduction of the viral load in children might act as a plausible mechanism by which BCG vaccination in children could reduce infections and mortality in a country.

Looking at an individual’s BCG vaccination history offers the possibility to perform better epidemiological studies than country-wide comparisons to determine vaccination schedules and strains that might optimize protection against COVID-19 as well as possible mechanisms of BCG protection. For instance, a recent study determined that BCG vaccination in childhood did not reduce COVID-19 infection in young adults, a population with reduced mortality due to COVID-19 (Hamiel, Kozer, and Youngster 2020). Patients with bladder cancer that have been treated with multiple doses of BCG (Alexandroff et al. 1999) constitute a population that is worth studying for possible protective effects against COVID-19.

If BCG were protective for COVID-19, why did COVID-19 spread in China despite having a universal BCG policy since the 1950s? During the Cultural Revolution (1966-1976), tuberculosis prevention and treatment agencies were disbanded and weakened(“Development and Expectation of Tuberculosis Service System in China” n.d.). We speculate that this could have created a pool of potential hosts that would be affected by and spread COVID-19. Currently, however, the situation in China seems to be improving.

Our data suggest that BCG vaccination is correlated with reduced mortality associated with COVID-19. However, there is still not proof that BCG inoculation at old age would boost defenses in elderly humans against COVID-19, but it seems to do so in Guinea pigs against *M. tuberculosis* (Komine-Aizawa et al. 2010).

BCG vaccination has been shown to produce broad protection against viral infections and sepsis(Moorlag et al. 2019), raising the possibility that the protective effect of BCG may not be directly related to actions on COVID-19 but associated co-occurring infections or sepsis. However, we also found that BCG vaccination correlated with a reduction in the number of COVID-19 reported infections in a country suggesting that BCG might confer some protection specifically against COVID-19. The broad use of the BCG vaccine across a population could reduce the number of carriers, and combined with other measures could act to slow down or stop the spread of COVID-19. However, an alternative explanation would be that COVID-19 in a person with BCG vaccination would have a milder presentation, reducing the possibility that such a case would be even detected in the first place.

Different countries use different BCG vaccination schedules(Zwerling et al. 2011), as well as different strains of the bacteria (Horwitz et al. 2009). We have not divided the data depending on the strain used to determine whether different strains are better at stopping the spread of infection, as well as reducing mortality in the elderly population, as suggested by Jun Sato(“If I Were North American/West European/Australian, I Would Take BCG Vaccination Now against the Novel Coronavirus Pandemic.” n.d.). As each country might have used the same strain for the whole population, the difference in strains for different purposes should be gathered in randomized control trials with different subjects from the same population tested with different strains.

Randomized controlled trials using BCG vaccination are required to determine how fast an immune response develops that protects against COVID-19. BCG is generally innocuous with the main side effect of inflammation at the site of injection. However, BCG is contraindicated in immunocompromised people as well as pregnant women(“Fact Sheets | Infection Control & Prevention | Fact Sheet - BCG Vaccine | TB | CDC” n.d.), so care should be taken when applying this possible intervention for COVID-19.

## Data Availability

Data used for the study is attached as supplementary material. All data used is publicly available.

## Acknowledgments

The authors would like to thank Raddy Ramos, Randy Stout, Isaac Kurtzer, Martin Gerdes, Kurt Amsler, Brian Harper, Madhukar Pai, Emily MacLean, Emma Risson and students, postdocs and faculty at the Immunology Institute of the Icahn School of Medicine, Mount Sinai for comments on the manuscript. We would like to thank Kashif Zafar for highlighting the widespread use of BCG vaccination by colonial powers in Asia and Africa between 1920s to 1950s, a critical piece of information.

## Notes

### Competing Interest Statement

The authors have declared no competing interest.

### Funding Statement

No external funding was received.

### Summary of Updates

-Updated updated data -Include European excess deaths analysis -Include multilinear analysis including population age, measures to control of epidemic, PBI per capita, health care conditions.

